# Pooled evidence precision of clinical trials on hydroxychloroquine for Covid-19 treatment was stabilized eight months after the outbreak

**DOI:** 10.1101/2024.01.21.24301572

**Authors:** Tatiane B Ribeiro, Paula C Ramirez, Luís Ricardo S Melo, Fredi A Diaz-Quijano

## Abstract

**OBJECTIVE:** At the beginning of 2020, hydroxychloroquine showed promising *in vitro* activity for Covid-19 and several studies were oriented to assess its safety and efficacy. However, after a few months, hydroxychloroquine has proved ineffective. The randomized controlled trials (RCTs) developed quickly and in different settings represent the scientific community’s capacity to assess drug repositioning effectiveness during a sanitary crisis. Therefore, a critical evaluation of the evidence generated can guide future efforts in analogous situations. We aimed to analyze the RCTs assessing the efficacy of hydroxychloroquine in treating Covid-19, describe their internal validity and power, and evaluate their contribution to the precision of the combined evidence for assessing the mortality outcome.

**STUDY DESIGN AND SETTINGS:** This meta-research included RCTs assessing hydroxychloroquine to treat patients diagnosed with Covid-19. It was part of an umbrella systematic review of methods/meta-research (PROSPERO: CRD42022360331) that included a comprehensive search in MEDLINE, EMBASE, Cochrane Library, and the Latin America Database - Lilacs. We retrieved studies published until January 10th, 2022. The risk of bias was assessed using Risk of Bias (RoB) 2.0. We analyzed methodology of the studies, precision and random error change through time from pooled evidence, study comparators, patient important outcome, power in different magnitude of effects proxy.

**RESULTS:** A total of 22 RCT were included, from that 17 (77%) assessed hospitalized patients and five (23%) outpatients setting. Mortality was related as primary endpoint in only 4 studies, however half of the studies included composite endpoints including mortality as a component. The internal validity analysis using RoB2 found that eight studies (36%) had a high risk of bias. Only one study had sufficient power to evaluate a moderate magnitude of effect (RR = 0,7 on mortality). The standard error to evaluate efficacy on mortality did not change appreciably after October 2020. From Oct 2020 to Dec 2021, 18 additional studies were published with 2,429 patients recruited.

**CONCLUSION:** This meta-research highlights the impact that collaborative, and network scientific research have on informing clinical decision-making. Duplicate efforts create research waste as precision analysis shows that after October 2020, there was not appreciably changes in the precision of the pooled RCT evidence to estimate the hydroxychloroquine effect on mortality.

**What is new?:**

- After Oct2020, grouped RCT on the use of hydroxychloroquine in Covid-19 showed that precision estimate has not been appreciably modified in subsequent studies.
- At least 18 RCT (n=2,429) could potentially be saved through collaborative work.
- Most individual studies did not have sufficient power to assess the size of moderate effect size on mortality.
- Strengthening cooperation and integrating research centers can decrease research waste.

## Introduction

Randomized controlled trials (RCTs) are considered the gold standard for identifying the causal effect between an intervention and a comparator, being the top primary evidence design in the hierarchy of evidence. However, the quality of a study is not only assessed by its design, but also by the rigor in conducting and analyzing the data (1). In addition, it’s important to consider other aspects of the body of evidence, such as the strength of association, risk of bias, imprecision, and indirect evidence (2).

During the Covid-19 outbreak, healthcare professionals had to critically assess the vast and emerging literature dynamically for clinical decision making (3). At that time, no Covid-19 vaccine was available, and it was urgent the fast and critical appraisal of the studies available for proper clinical decision. The best evidence available, mostly in the beginning of the pandemic, relied on observational studies, and only when robust RCTs were published did it become a game changer for Covid-19 treatment. Since early 2020, hydroxychloroquine (HCQ) demonstrated possibly promising activity for Covid-19 treatment and rapidly became widely used worldwide, driven by the media. Therefore, several RCTs and other studies were started during the pandemic. However, after a few months, hydroxychloroquine has proved ineffective (4). A critical evaluation of the evidence generated can guide future efforts in analogous situations. We aimed to analyze the RCTs assessing the efficacy of hydroxychloroquine in treating Covid-19, describe their internal validity and power, and evaluate their contribution to the precision of the combined evidence for assessing the mortality outcome.

## Methods

### General summary of the umbrella meta-research

This review was part of the umbrella systematic review/meta-research (PROSPERO registration: CRD42022360331) (5), that included studies that meet the following inclusion criteria patients diagnosed with COVID-19 using repositioning drugs (including hydroxychloroquine and chloroquine).

The methods were briefly described here, however full description can be found elsewhere. The review included a comprehensive search in MEDLINE (via PubMed), EMBASE, Cochrane Library, and the Latin America Database - Lilacs (via BVS). We retrieved studies published until January 10th, 2022, a period that included studies published during the Covid-19 sanitary emergency from early 2020 to the end of 2021. We searched for MESH terms and synonyms related to the intervention (repositioning drugs including hydroxychloroquine OR chloroquine) AND Covid-19 terms. Duplicates were removed, and pairs of reviewers screened independently all potential papers by titles and abstracts via Rayyan®. Discrepancies were resolved by consensus with a meeting with other researchers.

Studies that included “randomized” or “randomised” in their title or abstract had the following data extracted: general characteristics, outcomes data (mortality, mechanical ventilation, composite outcome: mortality plus mechanical ventilation, and primary data recorded by the study). In studies where the primary outcome was unclear or where more than one outcome was reported, we defined the outcome with the longest follow-up time or the greatest clinical importance (death) as the primary outcome. Finally, the risk of bias was assessed using Risk of Bias (RoB) 2.0 (6). Cross reference search included reference from studies included and WHO guideline plus main systematic reviews about the topic. (7; 4).All data extraction was performed by two independent reviewers, and discrepancies were resolved by consensus with a senior reviewer’s discussion.

### Eligibility criteria

We included randomized controlled trials that assessed hydroxychloroquine used as monotherapy, without combination, compared with placebo or standard of care, to treat patients diagnosed with Covid-19 (regardless of disease severity or setting).

### Data analysis

We analyzed population characteristics (hospitalized or outpatient, severity of disease), country recruitment, intervention, comparator, and primary outcomes, and overall risk of bias.

To assess the consistency between the evidence gathered, we carried out a meta-analysis of the RCTs to compare the summary results with previous systematic reviews (7;4). We also plotted in a historical timeline the most important facts related to hydroxychloroquine use during Covid-19 pandemic, with the novel information from RCT published.

The effect size was calculated considering three different scenarios:

- Very large effect size – Considering the GRADE framework: 0.2. Intervention 10% versus Control 50%. (8)
- Large effect size – Considering the GRADE framework: 0.5. Intervention 10% versus Control 20%. (8)
- Moderate/plausible effect size – Considering plausible effect sizes for interventions that had WHO recommendation for Covid-19, such as dexamethasone. Intervention 9.5% versus Control 13%. (7; 9)

A power value above 80% is often considered sufficient (10). Therefore, we reported the proportion of studies above 80% in each scenario (very large, large, and moderate/plausible effect size). We ran a metanalysis for mortality outcome, to obtain the standard error of logarithm of the relative risk (ln RR) with pooled data, after progressive chronological inclusion of each study. An additional analysis consisted in removing the Recovery and Solidarity trials, which were large sample-size studies. All data were analyzed using R 3.3.0.

## Results

From the 6,246 papers included in meta-research umbrella study, 146 papers were extracted as randomized controlled trials, from that 36 assessed HCQ (Figure 1). 17 studies were excluded: 6 studies used HCQ in association with other drugs in the intervention group (azithromycin, favipiravir, ritonavir-lopinavir, angiotensin receptor blockers), in 5 studies HCQ was the control, in 5 studies HCQ was stated as Standard of care (SoC) and in one RCT HCQ was found in both arms (aim to compare low vs high dose).

**Figure 1:**
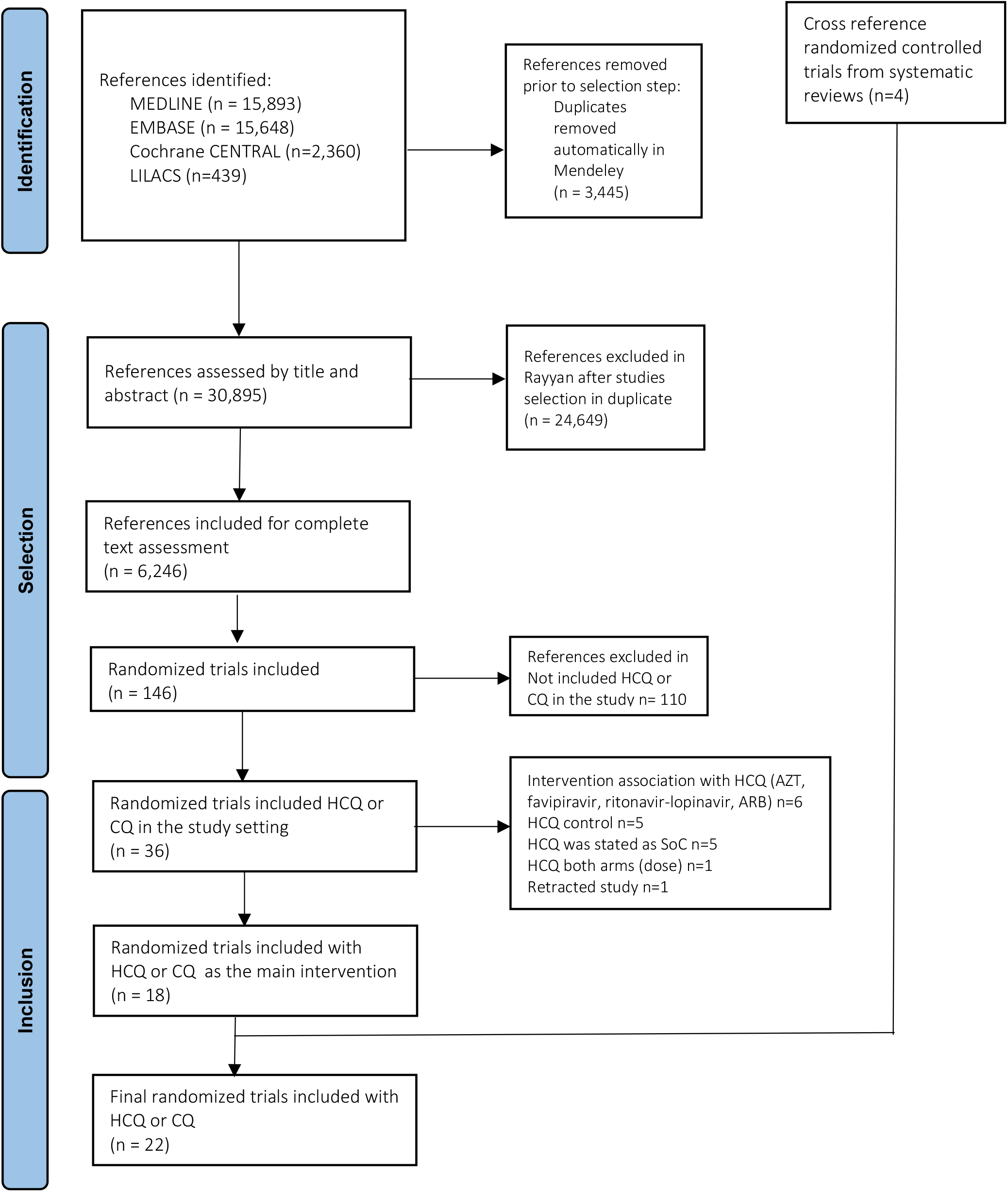
PRISMA flowchart of studies selection.

A total of 22 RCT (11; 12; 13; 14; 15; 16; 17; 18; 19; 20; 21; 22; 23; 24; 25; 26; 27; 28; 29; 30; 31; 32) that compared HCQ alone vs a comparator such as SoC, or placebo were included: 18 identified from the database and 4 from cross reference (Figure 1). Figure 2 presented the main historical facts related to hydroxychloroquine and chloroquine use since the beginning of the Covid-19 pandemic and included the main data from RCT published. Annex 1 from Supplementary material included the extracted data.

**Figure 2.**
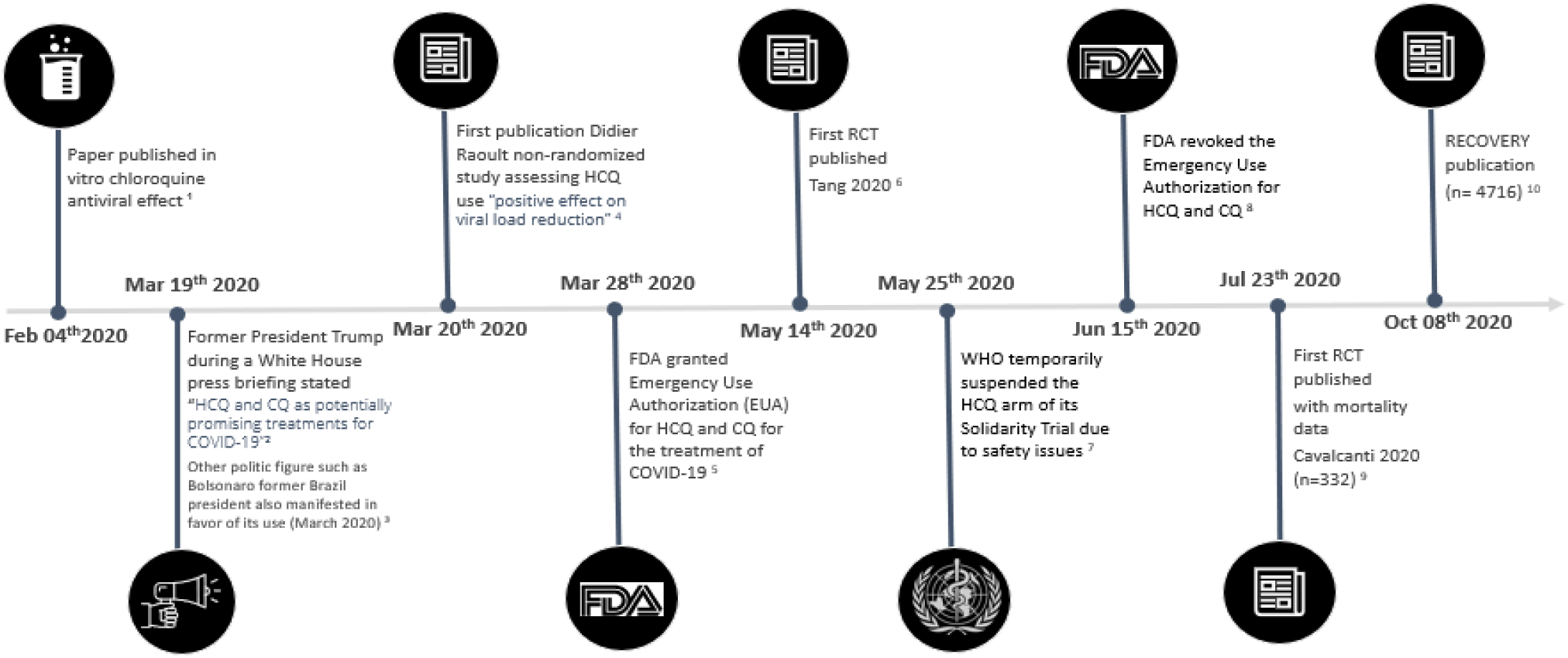
Historical timeline of hydroxychloroquine/chloroquine use during Covid-19 pandemic and spotlight RCT published. 1 Wang, M, et al. Remdesivir and chloroquine effectively inhibit the recently emerged novel coronavirus (2019-nCoV) in vitro. Cell Res 30, 269–271 (2020). https://doi.org/10.1038/s41422-020-0282-0 2 Trump’s history of promoting hydroxychloroquine. CNN. Available at: https://edition.cnn.com/videos/politics/2020/04/22/hydroxychloroquine-trump-covid-19-study-mh-orig.cnn [Access in Dec 10 2023] 3 Casarões, G., & Magalhães, D. (2021). A aliança da hidroxicloroquina: como líderes de extrema direita e pregadores da ciência alternativa se reuniram para promover uma droga milagrosa. Revista De Administração Pública, 55(1), 197–214. https://doi.org/10.1590/0034-761220200556 4 Gautret P, et al. Hydroxychloroquine and azithromycin as a treatment of COVID-19: results of an open-label non-randomized clinical trial. Int J Antimicrob Agents. 2020 Jul;56(1):105949. doi: 10.1016/j.ijantimicag.2020.105949. 5 Thomson, K., & Nachlis, H. (2020). Emergency use authorizations during the COVID-19 pandemic: lessons from hydroxychloroquine for vaccine authorization and approval. Jama, 324(13), 1282-1283. 6 Tang W, Cao Z, Han M, et al. Hydroxychloroquine in patients with mainly mild to moderate coronavirus disease 2019: open label, randomised controlled trial. BMJ 2020;369:m1849. doi: 10.1136/bmj.m1849 [published Online First: 20200514] 7 WHO temporarily suspended the HCQ arm of its Solidarity Trial, a global clinical trial testing potential treatments for COVID-19 (safety issues) 8 FDA News Release. Coronavirus (COVID-19) Update: FDA Revokes Emergency Use Authorization for Chloroquine and Hydroxychloroquine. Available in: https://www.fda.gov/news-events/press-announcements/coronavirus-covid-19-update-fda-revokes-emergency-use-authorization-chloroquine-and 9 Cavalcanti AB, Zampieri FG, Rosa RG, et al. Hydroxychloroquine with or without Azithromycin in Mild-to-Moderate Covid-19. N Engl J Med 2020;383(21):2041-52. doi: 10.1056/NEJMoa2019014 [published Online First: 20200723] 10 RECOVERY Collaborative Group; Horby P, et al. Effect of Hydroxychloroquine in Hospitalized Patients with Covid-19. N Engl J Med. 2020 Nov 19;383(21):2030-2040. doi: 10.1056/NEJMoa2022926. This article was published on October 8, 2020, at NEJM.org. Preliminary results posted on preprint server on July 15, 2020.

In the metanalysis of all the body of evidence the risk ratio was 1.08 (95% CI 0.99 to 1.08) (Supplementary material - Figure C), in line with a Cochrane systematic published previously (RR 1.09 - 95% CI 0.99 to 1.19) (4), and the BMJ living systematic review OR: 1.08 (IC 95% 0.92 to 1.27). (7) We found that 17 (77%) of the RCT published assessed hospitalized patients and five (22%) outpatients setting. Disease severity varied and only seven studies (32%) included severe Covid-19 patients (Table 1). Europe and North America were the regions with most publications, 6 (27%) and 5 (23%) respectively. Followed by South America, Africa, and Asia, four, three, and three respectively. Mortality the main patient important outcome, was related as primary endpoint in only four of studies, however half of the studies included composite outcome with mortality as a component endpoint. Six (27%) of the studies assessed laboratorial measures as primary endpoint (Table 1).

**Table 1.**
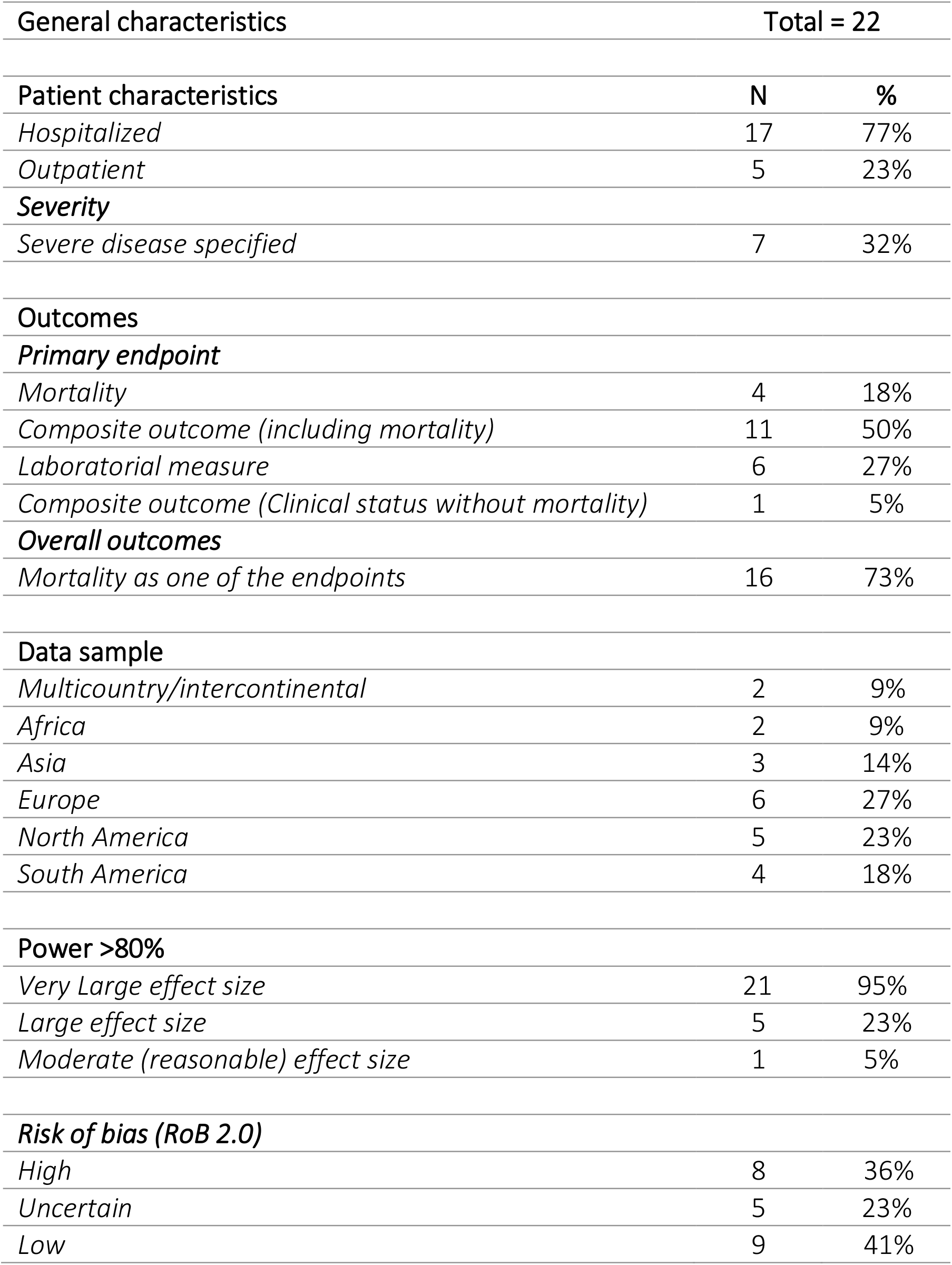
General characteristics of the included studies.

High risk of bias was seemed in eight studies (36%), most of them with limitation in the randomization process outcome (Figure 3). Nine studies had low risk of bias. Uncertain risk of bias was seen in 5 studies (23%).

**Figure 3:**
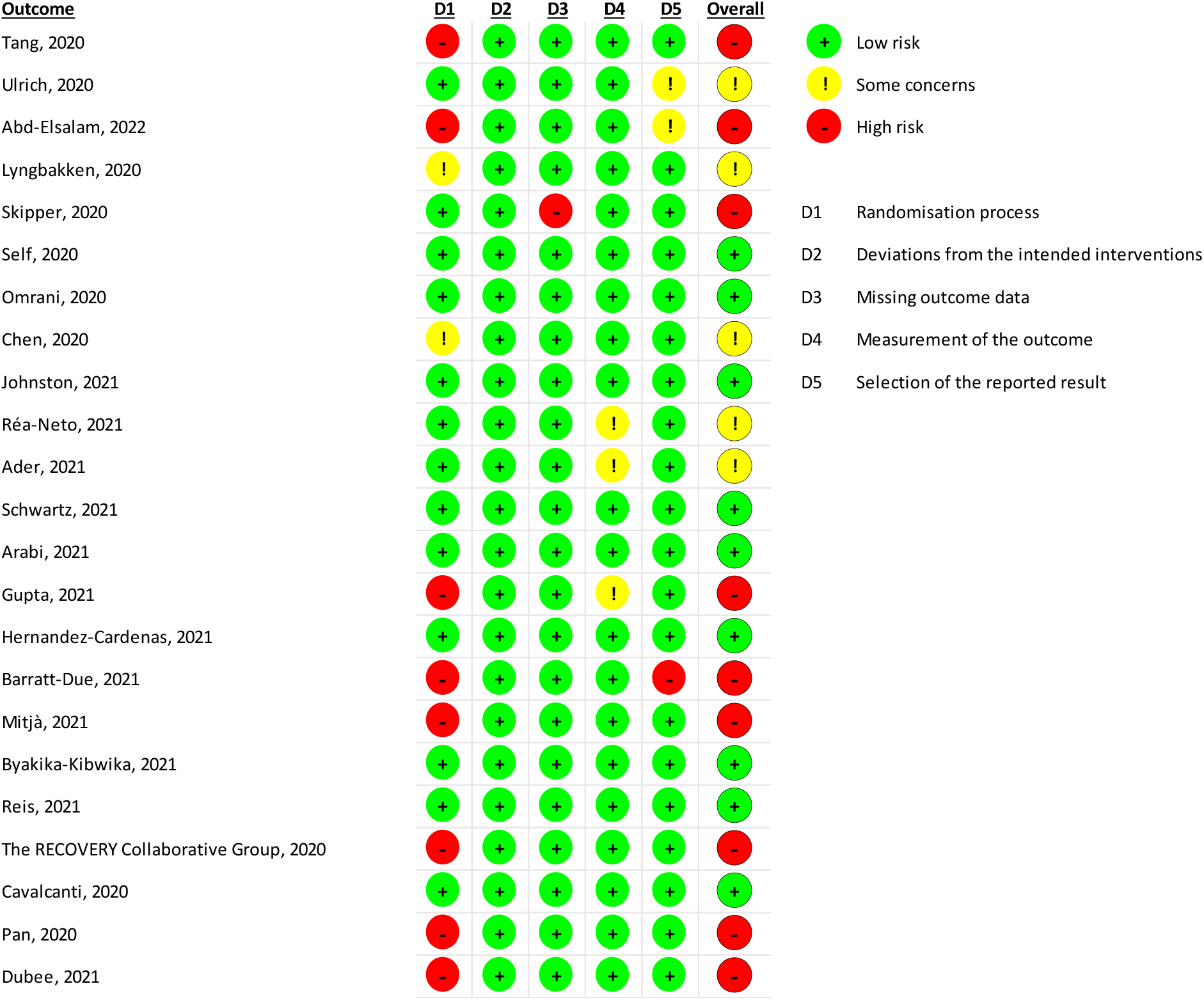
Risk of bias of included studies.

Only one study was powered enough (89%) to assess a moderate and reasonable magnitude of effect (RR = 0.7), and 5 studies could assess a large effect size (RR = 0.5). Most of the studies (21 of 22) had sufficient power to evaluate a very large magnitude of effect (RR = 0.2) (Table 1).

The precision of the treatment efficacy did not appreciably change with any new publication after October 2020 (Figure 4A – line blue). In the orange line from Figure 4A graph, we simulated the random error through time without considering the large RCTs (Recovery and Solidarity). Figure 4B showed the impact of including of new studies in sample size and number of events. After October 2020, 18 additional studies were published with 2,429 patients recruited.

**Figure 4A.**
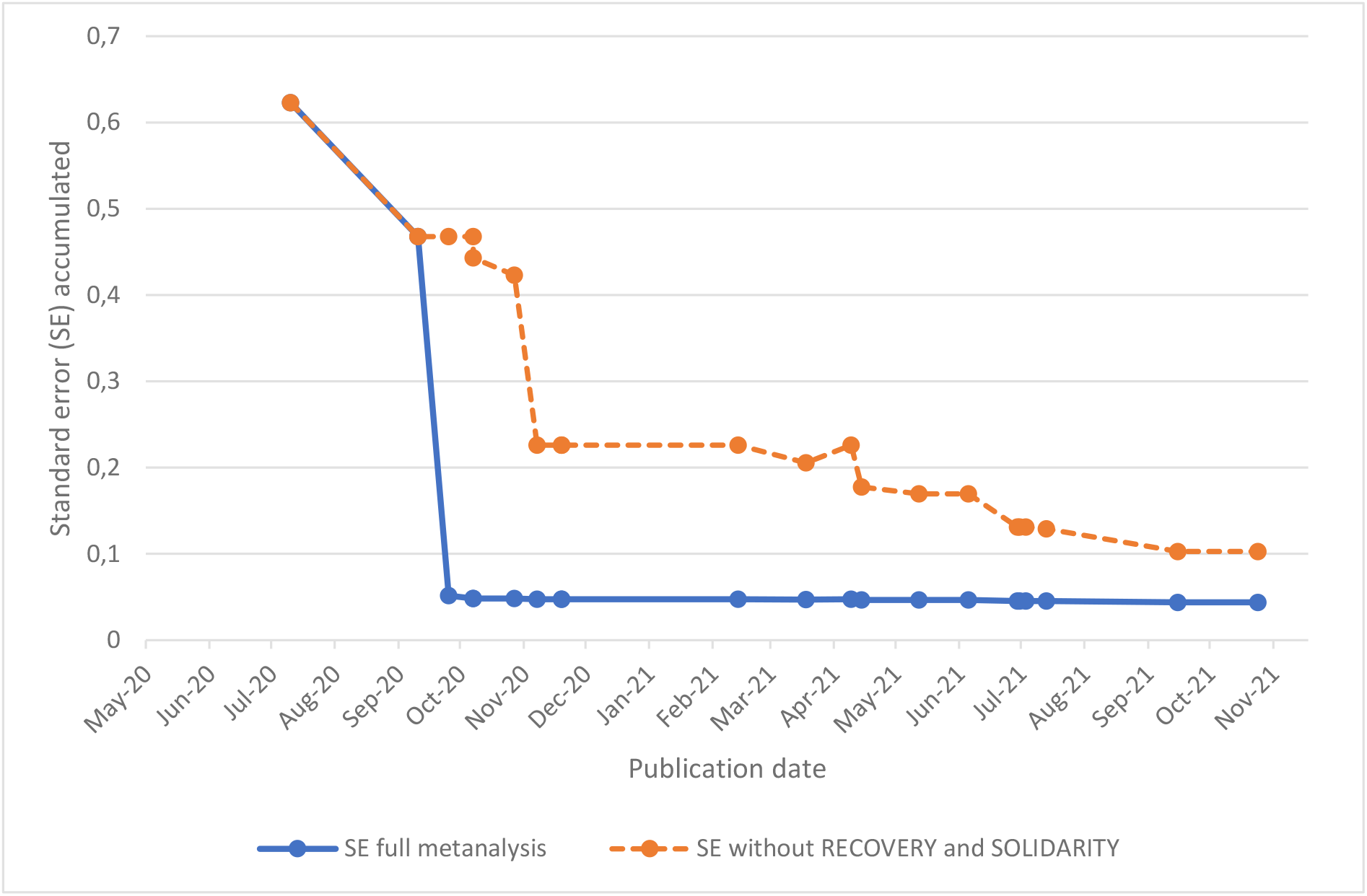
Effect of the chronological inclusion of new studies assessing mortality in the precision of summary of evidence at the time point.

**Figure 4B.**
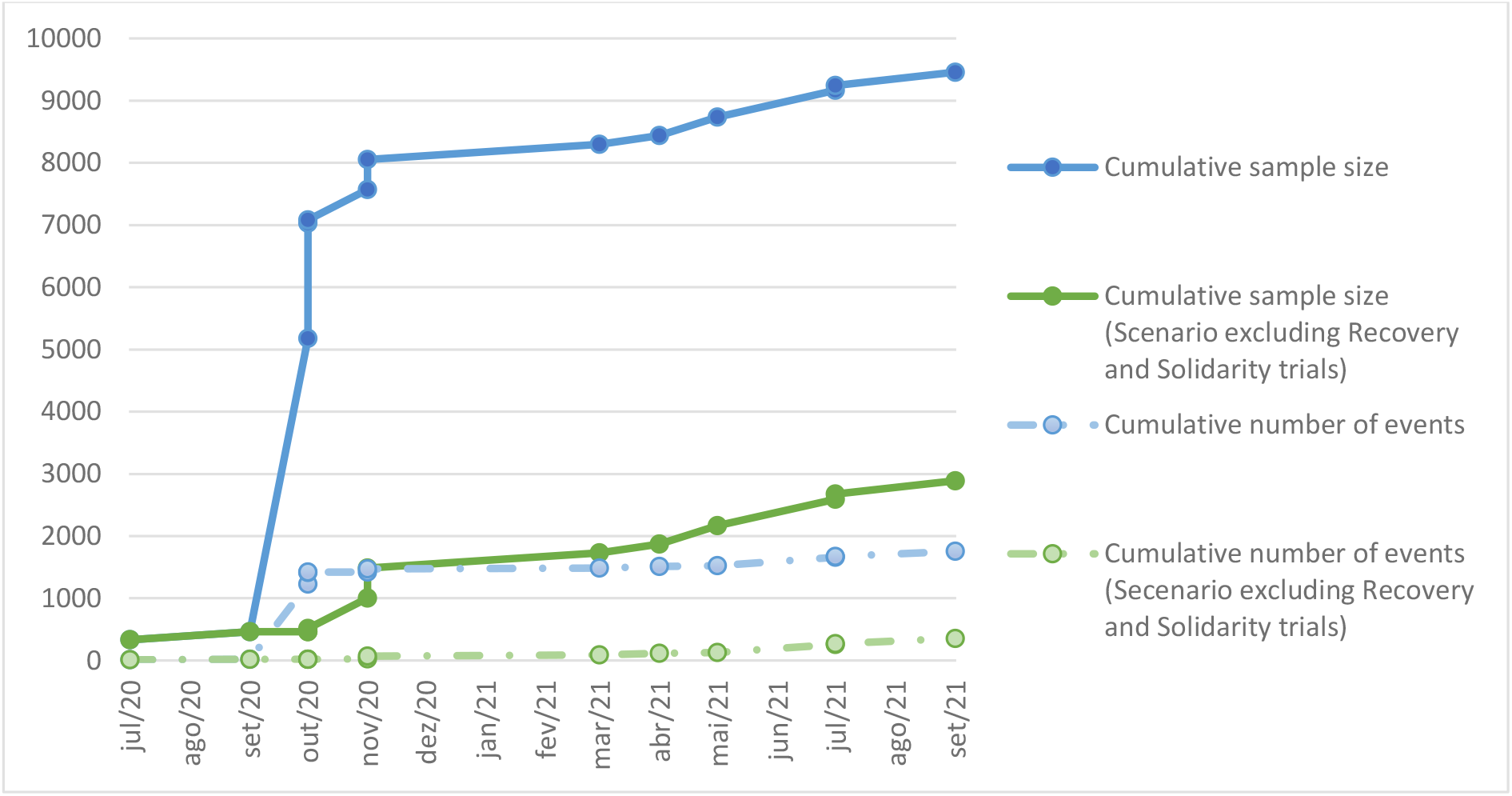
Effect of the chronological inclusion of new studies in the sample size and mortality events of summary of evidence at the time point.

## Discussion

Our paper found that eight months after the pandemic outbreak, the pooled evidence from RCTs resulted in a precision estimate of the mortality effect, suggesting that at least 18 RCT could potentially be saved through collaborative work. Recommendations on hospitals or by Healthcare Bodies around the world about compassionated use of hydroxychloroquine should be revised by October 2020.

The investment in time, money, and staff into the Covid-19 response reflects the worldwide humanitarian crisis. However, some efforts might be wasted, and focused research efforts were needed to proper response to evidence gaps, during the sanitary emergency. Research waste is a significant issue in the medical area. It has been estimated that up to 85% could be wasted due to poor research questions, study design, and unavailable reporting of results (33). In the Covid-19 era, these problems were amplified. Editorials by Glasziou, Sanders, and Hoffmann raised concerns about the number of protocols registered at ClinicalTrials.gov during the pandemic, with more than 1087 by May 2020 (34). Of these, 145 assessed hydroxychloroquine, suggesting duplicated research (34). Our study, which assessed all RCTs published in 2020 and 2021, confirmed this.

The research planning should include proper study designing and the choice of comparator is important for research validity (35). At early 2020, when it was unclear about the efficacy of hydroxychloroquine in Covid-19 treatment, the comparator in the scenario should be placebo or standard of care. In several studies, hydroxychloroquine was seen as standard of care, this analysis reflected just studies that clearly described it as standard of care in the comparator, however it known that other RCT included it as the drug was part of several hospitals protocol worldwide.

A previous review of protocols published in Brazil, in May 2020, described a large interest in hydroxychloroquine, with its use in combination and as a comparator (9). During the pandemic outbreak, many hospitals started using it relying on compassionate use, and at that time hydroxychloroquine was still being tested, and its effect was uncertain. For clinical matters, its use might be justified if a correct benefit-risk assessment is made individually. It is only possible to prove if a new intervention works after RCTs controlled with the current standard of care or placebo are published. A study published in Feb 2024 estimated that HCQ use was associated with an 11% increase in the mortality rate, by assessing observational studies published before reliable RCTs (36), highlighting the importance and need to generate quickly high-level evidence from RCTs in case of emergent diseases.

Another important element in preliminary study design is the appropriate outcome choice. We found that 74% reported mortality as one of the endpoints, but only 17% used mortality as the primary endpoint. The outcome choice needs to be relevant and important to key stakeholders, mainly patients (37). For Covid-19, patient-important clinical outcomes include mortality, respiratory failure, multiple organ failure, shortness of breath, and recovery. Research planning can include a consultation core outcome, for example in the COMET database, which is a comprehensive database with resources of Core Outcome Sets published following a validity methodology (e.g., Delphi method) (37).

The internal validity analysis using RoB2 found that eight studies (36%) had a high risk of bias, thus need to be assessed cautiously due to limited internal validity. This might also influence the certainty of pooled evidence (2) and change recommendations from clinical and health policy guidelines. Despite this, it is important to highlight some characteristics of the RoB2 tool that might penalize pragmatic trials, where commonly the concealment allocation might be challenging in multicenter.

The sample size choice is also part of the research planning process. We found that one study had sufficient power to identify a moderate magnitude of effect, while most of the studies would have power enough only to identify a very large magnitude of effect. It is known that a large magnitude of effect is rare in RCTs, mostly for death outcomes (38). Thus, study designs might be powered enough to demonstrate a reasonable effect in the real world.

According to the new revised pyramid, systematic reviews were the lens through which primary evidence was applied (1) to respond to a specific research question. The meta-analysis is the statistical aggregation that generate a single effect size (39; 1). Our study simulated how new evidence contribute to systematic reviews’ metanalysis, for responding to clinical questions that might change sanitary recommendation, as hydroxychloroquine use in Covid-19. In a scenario where we excluded larger sample size studies (Recovery and Solidarity), the standard error took much longer to decrease, and the clinical response remained uncertain for more time. Thus, studies well planned with collaborative network, such as Solidarity and Recovery, directly influenced clinical decision based on high certainty of evidence.

After the Covid-19 pandemic, researchers need to rethink research strategies. Strengthening cooperation and the integration of research centers will decrease research waste and increase power, value, and the use of efficient resources. Globalization research is necessary and more than expected in a fully connected world. The international collaborative multicenter clinical trials may shorten the timeline for clinical testing and decrease the cost of developing a new drug (40). Other novel initiatives to bring real-world questions addressing effectiveness included pragmatic trials and observational studies using big data might also be considered to leverage clinical research in emergency situations.

Our study included only published papers, and we know that some of the recommendations for Covid-19 relied on preprints, which did not always turn into peer-reviewed papers. Despite it, our meta-analysis had similar data from other published systematic reviews, suggesting that the studies included were representative of the evidence available in the literature.

This meta-research highlights the importance of collaborative and network scientific research in informing clinical decision-making, especially during sanitary emergencies. Agile knowledge generation is crucial for patient care, and it should be aligned with efforts to strengthen cooperation and integration among research centers to reduce waste and increase research value.

## Data Availability

All data produced are available online as all studies included were published

## SUPPLEMENTARY MATERIAL

**Table 1:**
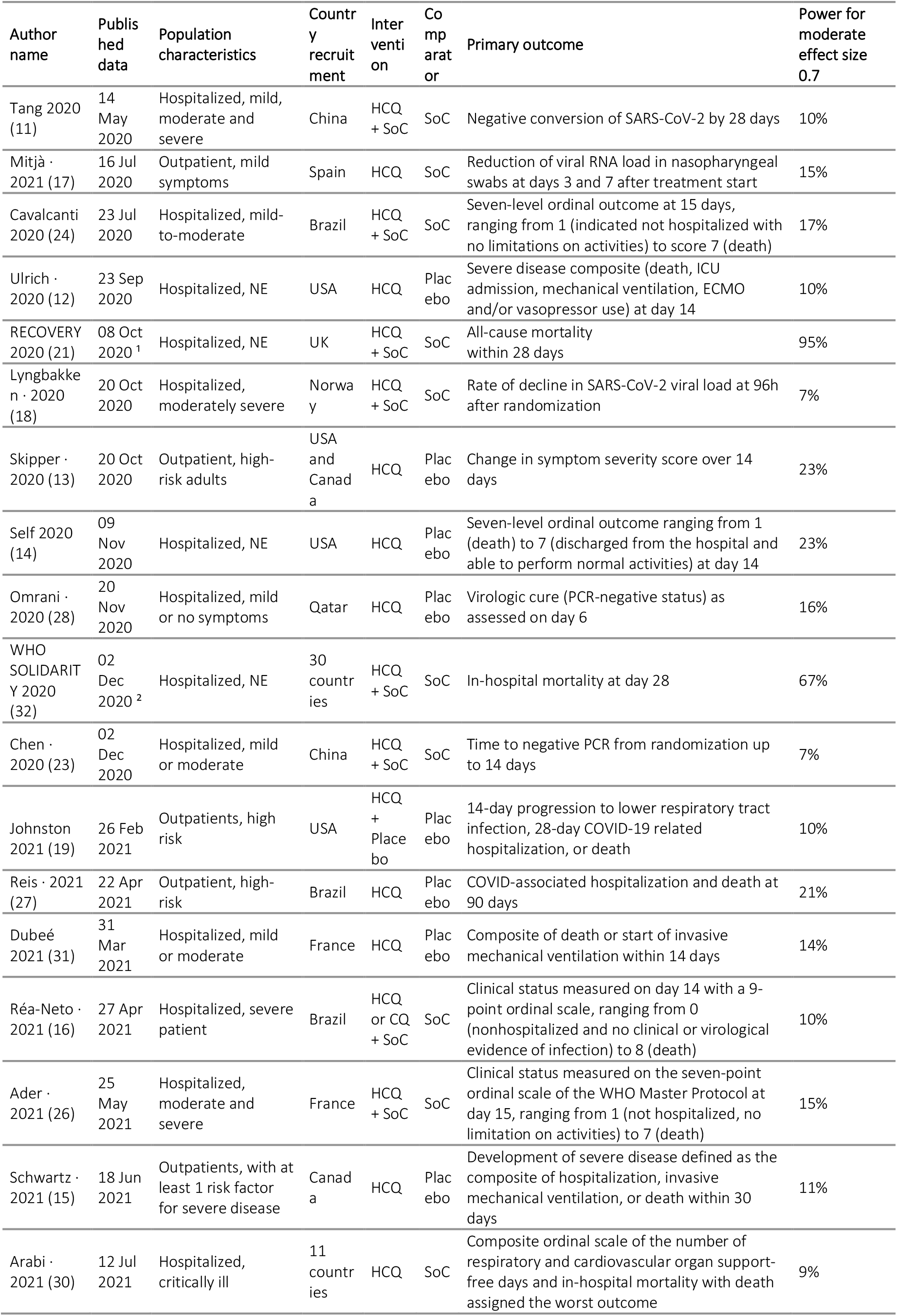
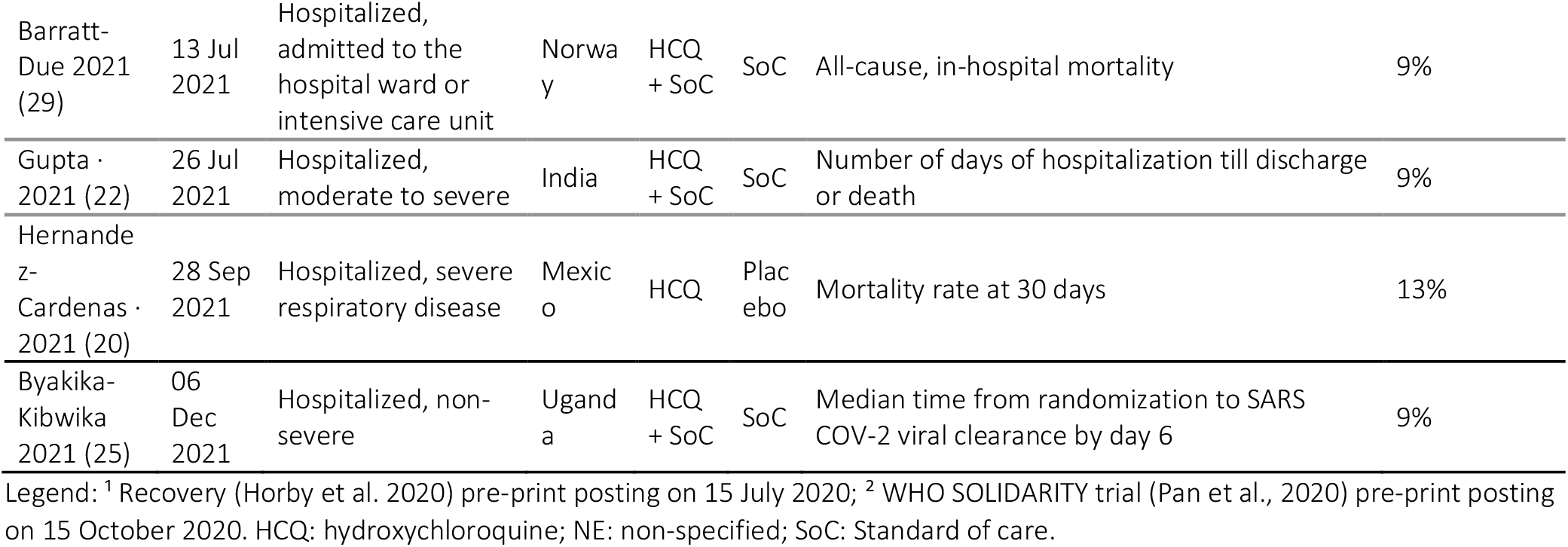
Description of the 22 RCT assessing HCQ efficacy.

**Figure B:**
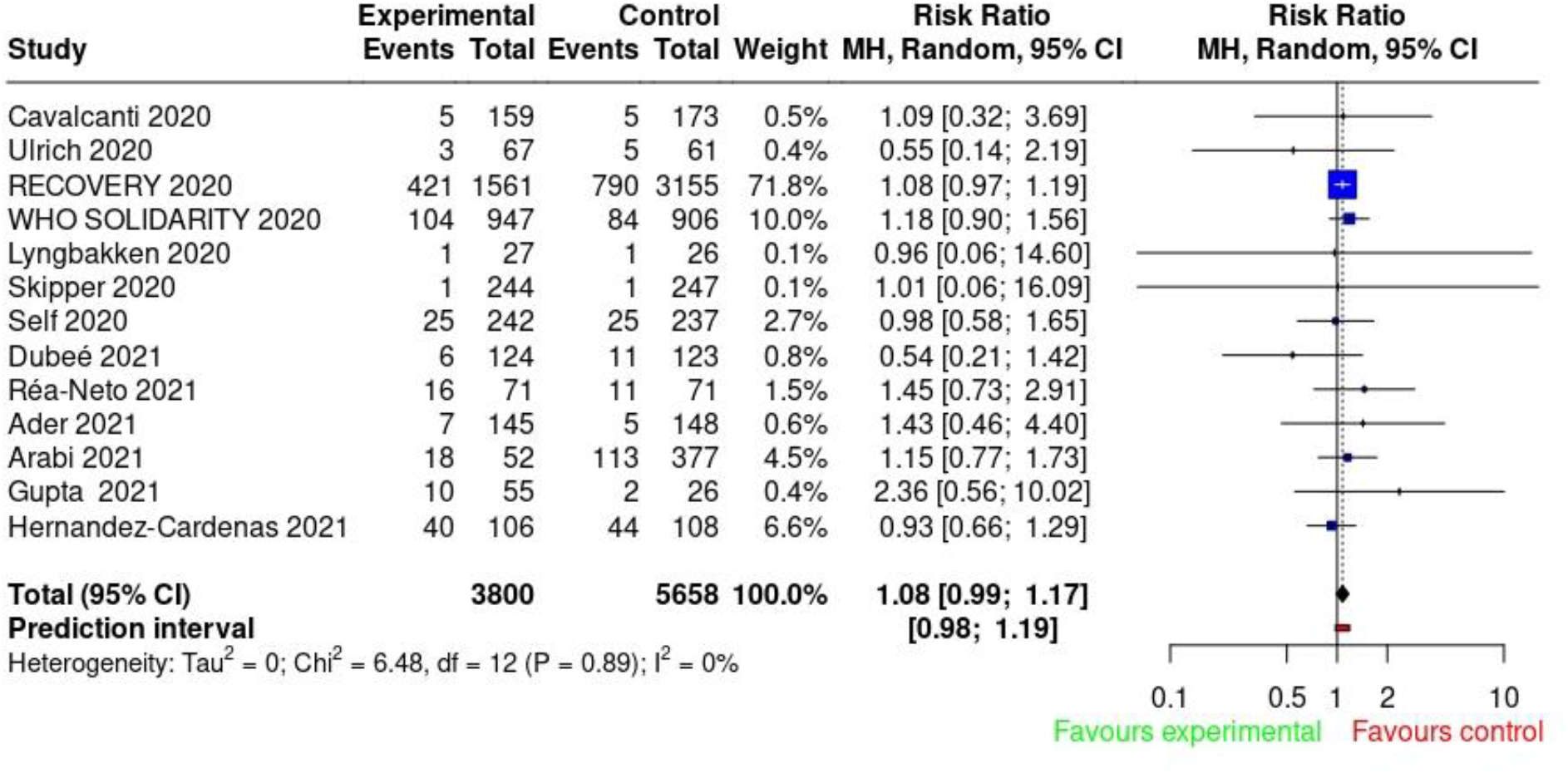
Forest plot for studies assessing mortality.

